# Does higher BMI increase COVID-19 severity?: a systematic review and meta-analysis

**DOI:** 10.1101/2020.12.05.20244566

**Authors:** Akibul Islam Chowdhury, Md. Fazley Rabbi, Tanjina Rahman, Sompa Reza, Mohammad Rahanur Alam

**Affiliations:** Department of Food Technology and Nutrition Science, Noakhali Science and Technology University, Bangladesh; Department of Biotechnology and Genetic Engineering, Noakhali Science and Technology University, Bangladesh; Institute of Nutrition and Food Science, University of Dhaka, Bangladesh

**Keywords:** COVID-19, Overweight, Obesity, BMI

## Abstract

**Introduction:** COVID-19 pandemic has caused havoc worldwide, and different comorbidities have been seen to exacerbate the condition. Obesity is one of the leading comorbidities, which is associated with many other diseases. In this paper, we present a systematic review and meta-analysis estimating the effects of overweight and obesity on COVID-19 disease severity.

**Methodology:** Two electronic databases (Medline and Cochrane library) and one grey literature database (Grey Literature Report) were searched using the following keywords: overweight, obesity, body mass index, respiratory disease, coronavirus, COVID-19. The risks of bias of the selected studies were assessed by using the Navigation Guide method for human data. Both random and fixed effect meta-analysis were determined using Review Manager (RevMan) software version 5.4.

**Results:** After initial screening, 12 studies (7 cohort studies, four case-control studies, and one cross-sectional study) were fulfilled the eligibility criteria, comprising a total of 405359 patients and included in the systematic review. The pooled risk of disease severity was 1.31 times higher based on both fixed and random effect model among those overweight patients, *I*^*2*^ 0% and 2.09 and 2.41 times higher based on fixed and random effect respectively among obese patients, *I*^*2*^ 42% compared to healthy individuals.

**Conclusion:** Overweight and obesity are common risk factors for disease severity of COVID-19 patients. However, further assessment of metabolic parameters included BMI, waist-hip ratio, and insulin levels, are required to estimate the risk factors of COVID-19 patients and understanding the mechanism between COVID-19 and body mass index.

## Introduction

Coronavirus disease 2019 (COVID-2019)—caused by the severe acute respiratory syndrome coronavirus 2 (SARS-CoV-2) virus—was declared a pandemic by the World Health Organization on March 11, 2020 (1). As of December 1, 2020, COVID-19 has infected 61.8 million people worldwide, with a death toll of 1.4 million (2). Previously, two highly pathogenic Coronaviruses resulted in outbreaks of a severe acute respiratory syndrome (SARS) in 2003 in Guangdong province, China, and the Middle East respiratory syndrome (MERS) in Middle Eastern countries in 2012 (3-6). Multiple risk factors are associated with mortality in COVID-19 patients. An increasing body of data suggests that individuals with diabetes mellitus (7), hypertension, and severe obesity (BMI ≥ 40 kg/m^2^) are more likely to be infected and are at a higher risk for complications and death from COVID-19 (8-14). In China, it was found that patients with cardiovascular disease (10.5%) had the highest fatality rate than diabetes mellitus (7.3%), hypertension (6.0%), and cancer (5.6%) (15). Many countries mentioned body mass index (BMI) as a clinical risk factor of COVID-19, such as China (16), Italy (17), United States (9) as the immunity system plays a vital role in obesity-induced adipose tissue inflammation (11). Emerging literature suggests that adults with obesity under the age of 60 are more likely to be hospitalized (18). The prevalence of obesity among adults is increasing day by day due to insufficient physical activities. A previous study showed a strong correlation between obesity and complications of viral infections (influenza virus, SARS, and MERS) (19). Many studies found that excessive weight gain ≥18 kg may increase the risk of developing community-acquired pneumonia (7, 20). Severe obesity might increase the duration of hospital stay and the case fatality rate (14, 21). However, two earlier reports have suggested no difference in body mass index (BMI) between severe and non-severe groups (18, 22). Although several studies addressed the impact of the body mass index (BMI) on COVID□19, a definite conclusion has not been drawn yet. Hence, this meta-analysis was conducted to elucidate the relationship between obesity and COVID-19 by searching existing literature.

## Methods

### Literature Search

We searched two electronic databases: MEDLINE (on October 20, 2020) and Cochrane library (on October 21, 2020). We also searched one grey literature database: Grey Literature Report (http://greylit.org) on October 21, 2020. Searches were performed using a search strategy in English. We searched the literature using the following keywords: overweight, obesity, body mass index, respiratory disease, coronavirus, COVID-19. Manual searching was also performed to identify potentially eligible studies.

### Study selection

All articles found in the searches were downloaded, and duplicate articles were identified and excluded. Two independent authors screened the titles and abstracts for finding duplicates and then screened the full texts to select the eligible articles. If there were any disagreements between the review authors, a third author fixed the problems. Following the PRISMA guideline, the process of study selection is presented in a flow chart (Figure 1).

**Figure 1:**
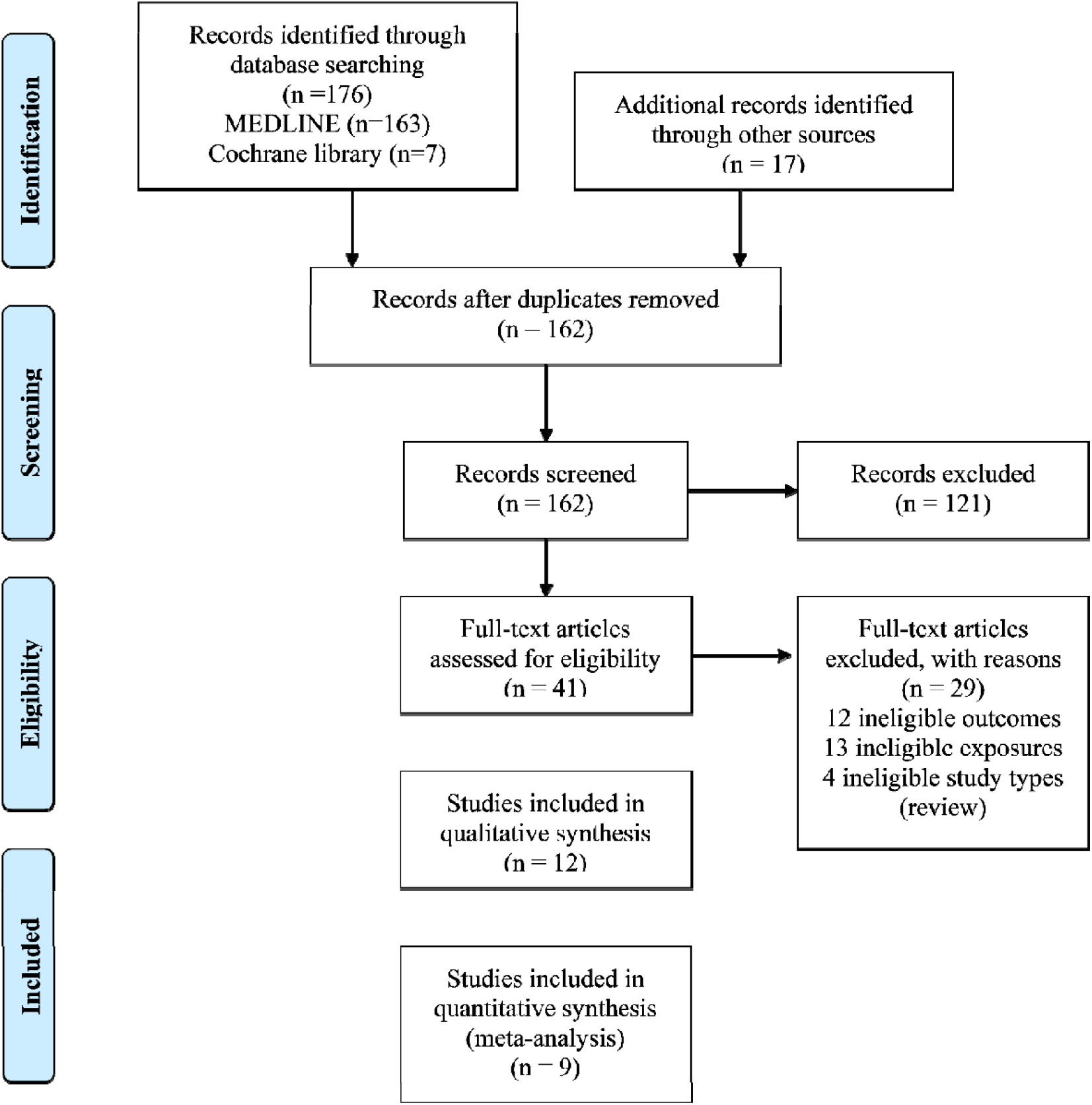
The flow chart of searching and selecting studies based on selected criteria for systematic review and meta-analysis.

### Eligibility criteria

PECO definitions are described below:

- ***Population***: We included all studies of people aged (≥ 15 years) and reported positive for the presence of coronavirus in their bodies by the RT-PCR technique. Studies that measured the body mass index (BMI) were included with standard procedure.
- ***Exposures:*** Studies that defined overweight and obesity with standard definition were included.
- ***Comparators:*** Healthy participants with optimum BMI were used as a comparator. All other comparators were excluded.
- ***Outcomes:*** Severity of disease was used as an outcome in this systematic review.

### Types of study

We included studies that measured the effect of overweight and obesity on COVID-19 disease severity. Eligible studies were randomized control trials, cohort studies (both prospective and retrospective), and case-control studies. We also included observational study. Records published in English were included. Both published and unpublished studies were included. Studies conducted using unethical practices were excluded.

### Types of effect measures

We included measures of the relative effect of overweight and obesity on the severity of disease (prevalence and incidence), compared with the patient with optimum BMI. We included relative effect measures such as RRs, ORs, and Hazard ratios. To facilitate meta-analysis, if a study presented an RR, then it was converted to OR. If a study presented estimates for effect from two or more alternative models that had been adjusted for different variables, then we systematically prioritized the estimate from the model that provided information on the relevant confounders or mediators, at least the core variables: age, sex, and socioeconomic position. We prioritized estimates from models adjusted for more potential confounders over those from models adjusted for fewer. For example, if a study presents estimates from a crude, unadjusted model (Model A), a model adjusted for one potential confounder (e.g., age; Model B) and a model adjusted for two potential confounders (e.g., age and sex; Model C), then we prioritized the estimate from Model C.

### Data extraction

Two independent reviewers extracted the data on study characters (study authors, study country, population size, study year, exposure, and outcome), study design, and risk of bias (including source population representation, blinding, exposure assessment, outcome assessment, confounding, incomplete outcome data, selective outcome reporting, conflict of interest and other sources of bias).

### Risk of bias assessment

There is no standard method of assessing the risk of bias of selected studies for doing systematic review. The risk of bias of this review was assessed by nine risk factors of bias included in the Navigation Guide method for human data. These were: (i) source population representation; (ii) blinding; (iii) exposure assessment; (iv) outcome assessment; (v) confounding; (vi) incomplete outcome data; (vii) selective outcome reporting; (viii) conflict of interest; and (23) other sources of bias. The ratings for all domains were: “low”; “unclear” and “high.” Two independent reviewers assessed the risk of bias of selected studies. Any disagreement was solved by the third reviewer. Funnel plots were generated to judge concerns on publication bias (Supplementary figures).

### Statistical Analysis

We assessed heterogeneity by reporting the *I*^2^ (% residual variation due to heterogeneity) and tau^2^ (method of moments estimates of between-study variance) of the pooled estimate. Both random effect and fixed-effect models were used to measure the relationship between obesity and COVID-19 disease severity. The 95% confidence interval has been reported in a pooled analysis. All analysis was done by using Review Manager (RevMan) software version 5.4.

## Result

### Study selection

A total of 193 individual studies were identified in our searches. Twelve studies fulfilled our eligibility criteria and were included in the systematic review (Figure 1). Of the 12 included studies (24-35), nine studies were included in the meta-analysis (24, 26-30, 32, 34, 35).

### Characteristics of included studies

Most of the studies were cohort studies (7 studies), followed by case-control studies (4 studies) and one cross-sectional analysis. The total population of the included studies was 405359. The most commonly studied countries were the United States (6 studies) and China (4 studies). The comparator of most studied was BMI≤ 25kg/m^2^ (Table 1).

**Table 1:**
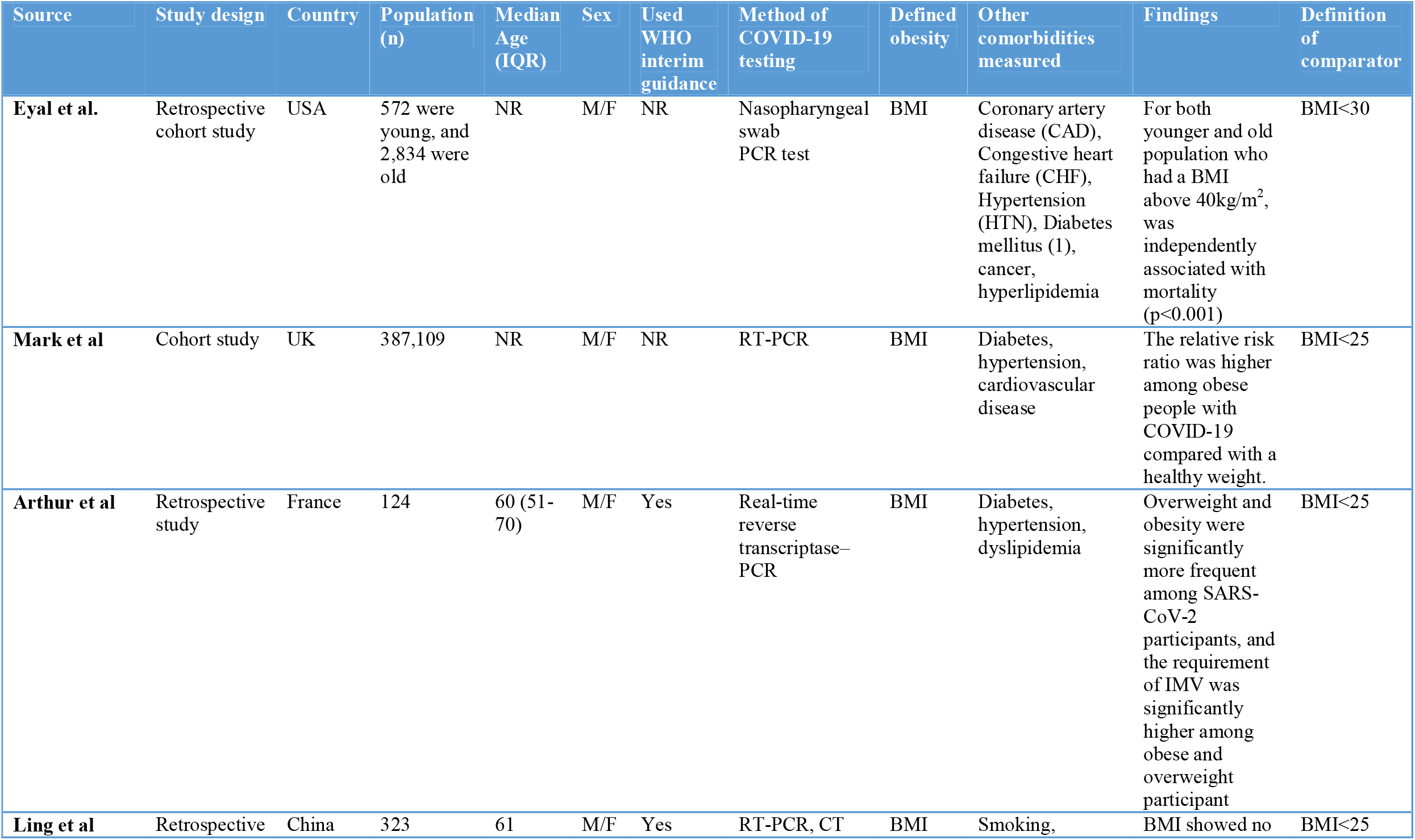
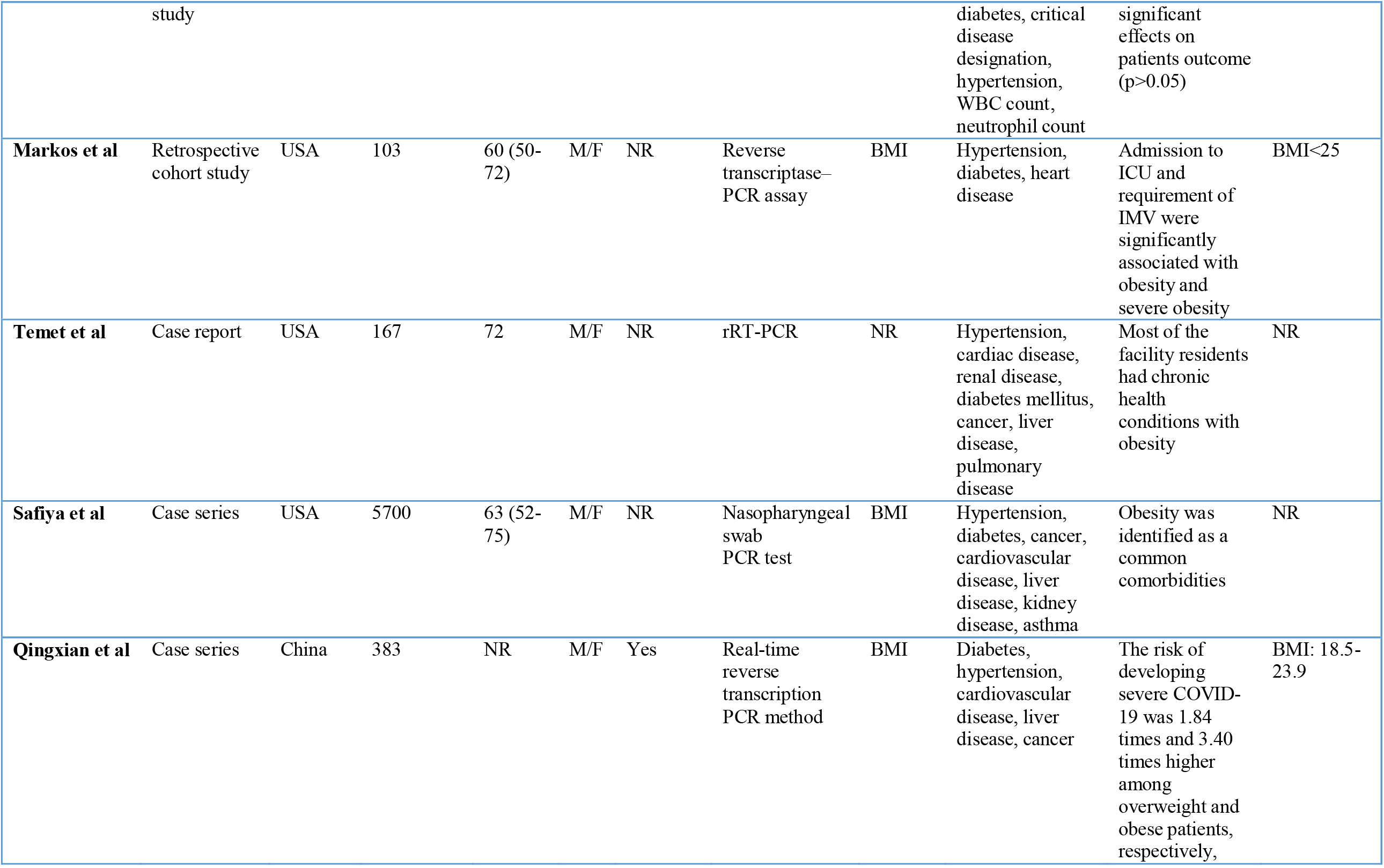
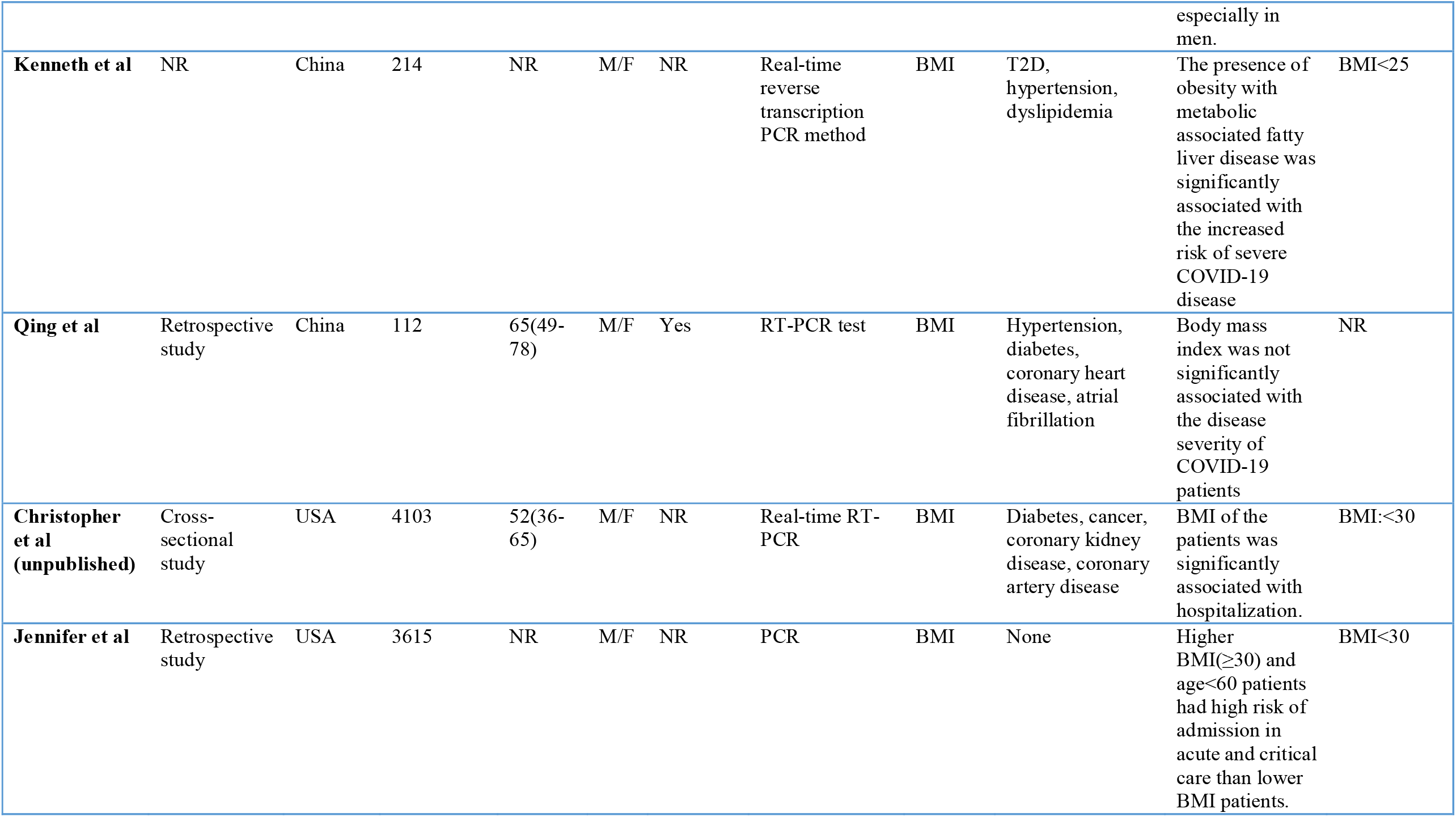
Result of systematic review (published and grey article)

Nine studies reported the relation between COVID-19 disease severity with overweight and obesity (Table 3).

**Table 3:**
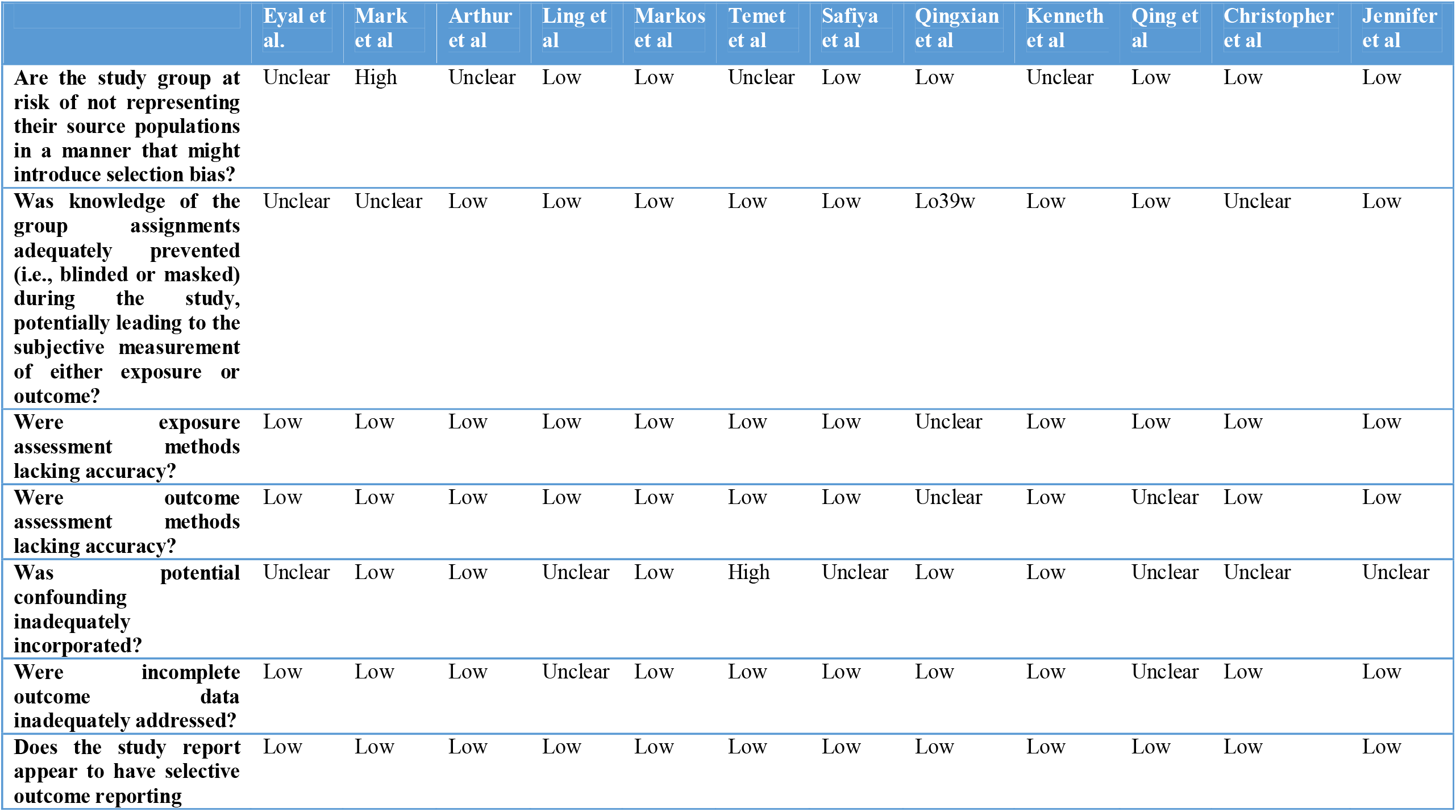
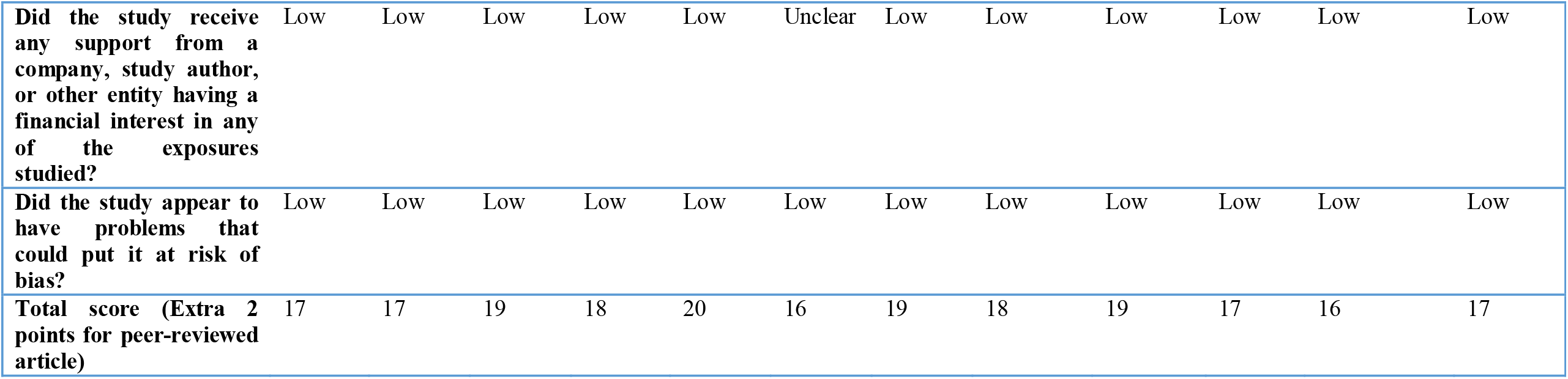
Risk of bias assessment.

**Table 3:** Odds ratio of selective studies for meta-analysis.

|  | Eyal et al | Arthur et al | Markos et al | Kenneth et al | Ling et al | Qingxian et al | Mark et al | Jennifer et al | Christopher et al |
| --- | --- | --- | --- | --- | --- | --- | --- | --- | --- |
| **Overweight** | 1.1(0.5-2.3) | 1.69(0.52-5.48) | 2.27(0.59-8.83) | --- | 0.65(0.19-2.23) | 1.74(1.03-2.93) | 1.32(1.09-1.60) | 1.1(0.8-1.7) | 1.38(1.03-1.85) |
| **Obese** | 5.1(2.3-11.1) | 7.36(1.63-33.14) | 5.39(1.13-25.64) | 6.32(1.16-34.54) | 2.86(0.79-10.31) | 2.69(1.31-5.52) | 1.97(1.61-2.42) | 1.5(0.9-2.3) | 1.73(1.03-2.90) |

**\*Reference: Person with a normal BMI (18.5-24.9 wt/m<sup>2</sup>)**

### Risk of bias at individual study level

The risks of bias rating for each domain for all 12 studies for this outcome are presented in Table 2.

### Measured outcome

The effect of overweight on disease severity of COVID-19 patients was measured comparing with normal body weight. The meta-analysis of selected nine studies showed that the pooled risk of disease severity was 1.31 times higher based on both fixed and random effect model among those patients who were overweight, *I*^*2*^ 0% (Figure 2 and 3).

**Figure 2:**
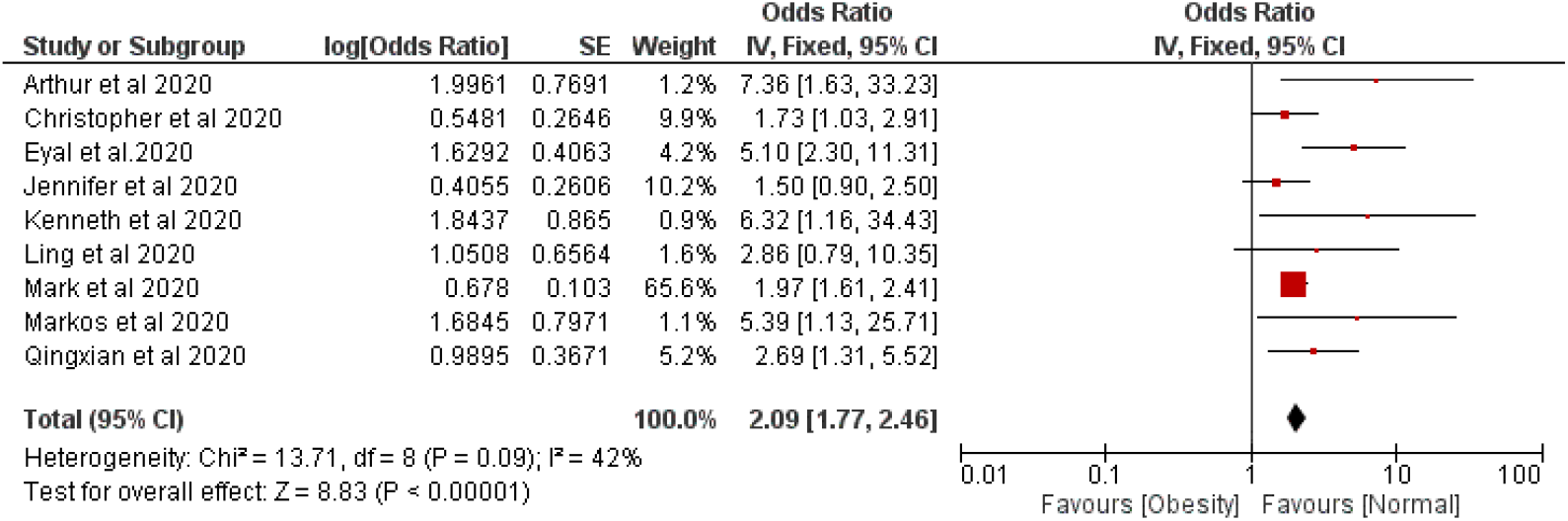
Forest plot illustrating the Fixed effect model of the association between obesity and COVID-19 severity.

**Figure 3:**
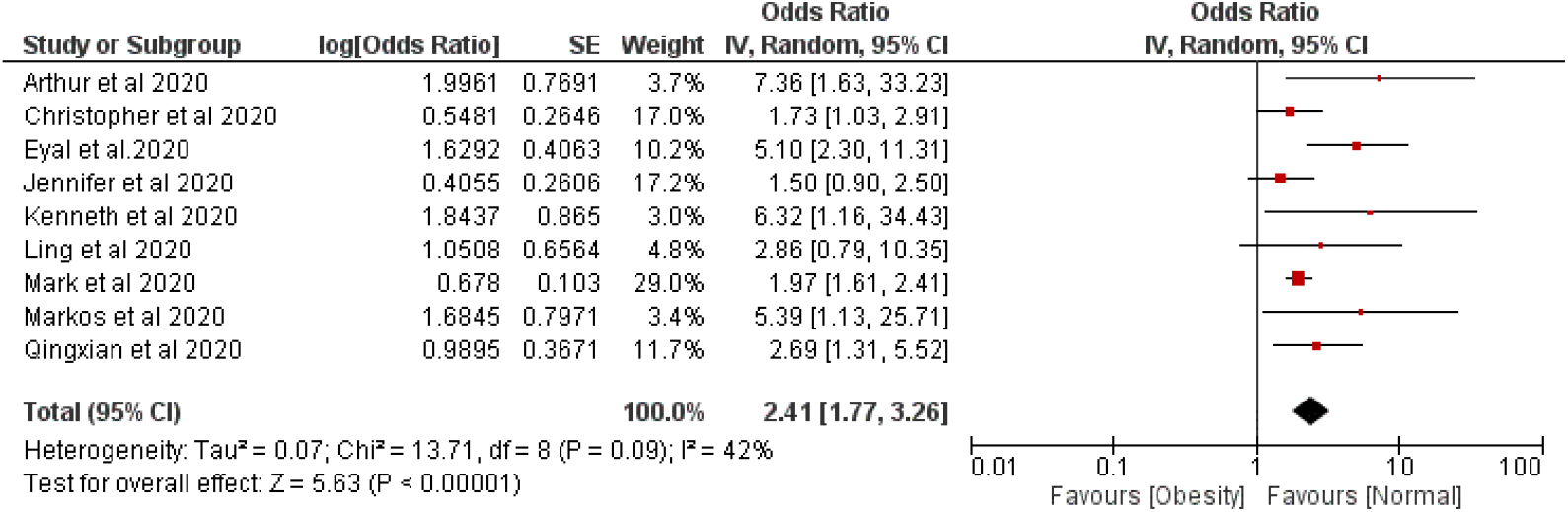
Forest plot illustrating the Random effect model of the association between obesity and COVID-19 severity.

The pooled risk of disease severity was 2.09 and 2.41 higher based on fixed and random effect respectively among obese patients compared with normal body weight patients, *I*^*2*^ 42% (Figure 4 and 5).

**Figure 4:**
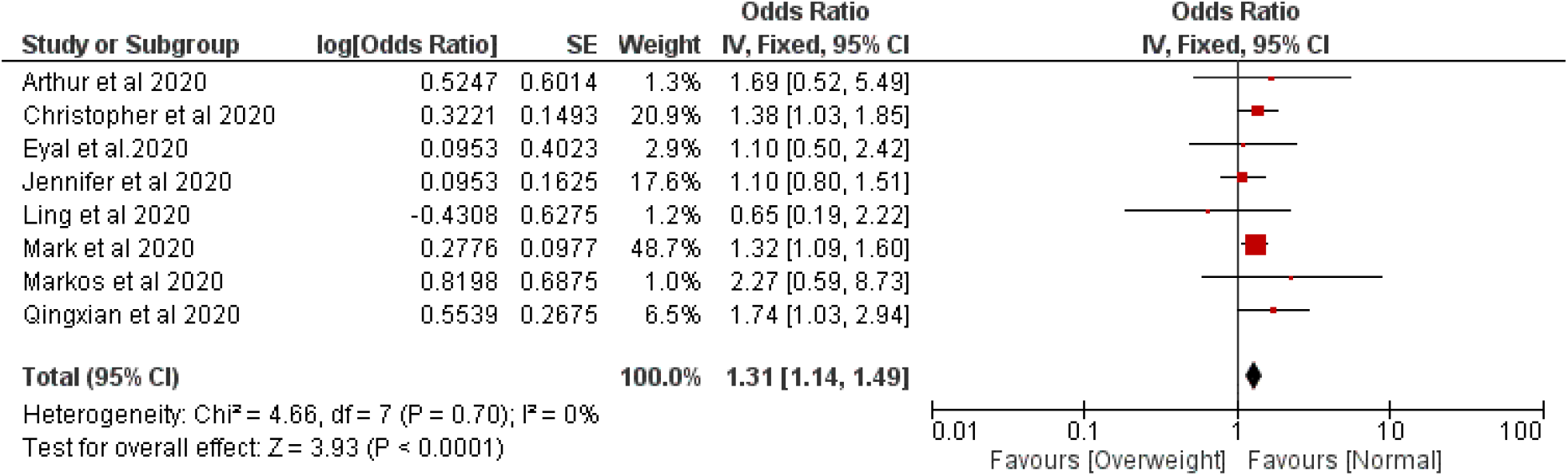
Forest plot illustrating the Fixed effect model of the association between overweight and COVID-19 severity.

**Figure 5:**
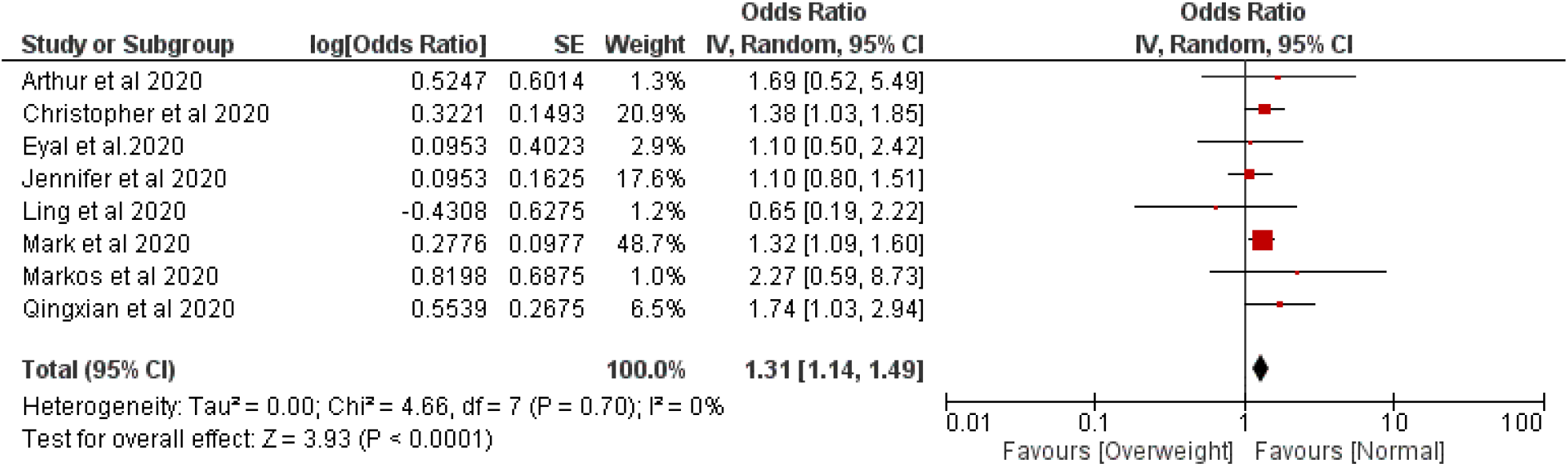
Forest plot illustrating the Random effect model of the association between overweight and COVID-19 severity.

## Discussion

Of the 41 studies examined, some studies failed to find any association between BMI and COVID-19 (25, 27), and some studies did not measure BMI as a risk factor of COVID-19. The present study accumulated all the findings related to COVID-19 and BMI. Interestingly, it was found that BMI is a risk factor for COVID-19 patients. Overweight patients were 1.31 times higher at the risk of disease severity of COVID-19, and obese patients were 2.09 and 2.41 times higher susceptible to the severity of COVID-19 according to fixed and random effect model, respectively. Our study result is consistent with what has been found in previous reports (36-40). Different studies previously documented for different viral pathogens, including influenza, that obesity was a major risk factor for disease severity (41-43). During the 2009 H1N1 pandemic, it was found that the rates of hospitalization and deaths were higher among overweight and obese patients (7).

Several parameters with overweight and obesity play a role in disease severity of COVID-19. However, there is no exact mechanism that explains the contribution of overweight and obesity to severe COVID-19 outcomes. Nevertheless, it is said that obesity has adverse effects on lung function, diminishing forced expiratory volume and forced vital capacity (44). It is also reported that respiratory physiology is changed by obesity with the decreased functional capacity of the respiratory system (45). Another study found that obesity impaired immune system surveillance and response (46). Obesity was also found to impair the respiratory function, gas exchange, lung volume, increase comorbidities (CVD, T2D, kidney disease), and metabolic risk (hypertension, insulin resistance, and dyslipidemia), which contributed to disease severity of COVID-19 patients (47). Some studies explained why obese people presented a worse clinical outcome than a normal patient. These studies concluded that overweight and obese people have a different innate and adaptive immune response and have higher leptin and lower adiponectin concentrations, which leads to dysregulation of immune response and contributes to worsening pathogenesis conditions (48-50). Another study found that obesity reduced the activity of macrophages when an antigen is presented (51). Obesity was also directly associated with basal inflammatory status characterized by higher circulating Interleukin 6 and C-reactive protein levels (44). Obesity also impaired the adaptive immune system responses to the influenza virus (52). It is crucial to understand and find out the relationship between obesity and COVID-19 to reduce the risk of developing severe COVID-19 illness. The lifestyle of people should be improved to lessen risk both in the current and subsequent waves of COVID-19.

## Conclusion

Further assessment of metabolic parameters, including BMI, waist-hip ratio, and insulin levels, are required to estimate the risk factors of COVID-19 patients. Obesity is a common risk factor of numerous comorbidities that increase the severity of COVID-19 disease. So, we recommend that additional attention should be given to obese patients, along with other patients during this epidemic.

## Supporting information

Funnel plots were generated to judge concerns on publication bias (Supplementary figures).

Funnel plots were generated to judge concerns on publication bias (Supplementary figures).

Funnel plots were generated to judge concerns on publication bias (Supplementary figures).

Funnel plots were generated to judge concerns on publication bias (Supplementary figures).

## Data Availability

The datasets generated during this study are available from the corresponding author on a reasonable request.

## Acknowledgment

We would like to express our gratitude to the authors of the studies included in our study.

