## Supplementary figures and images for "Does higher BMI increase COVID-19 severity?: a systematic review and meta-analysis"

### Funnel plots were generated to judge concerns on publication bias (Supplementary figures).

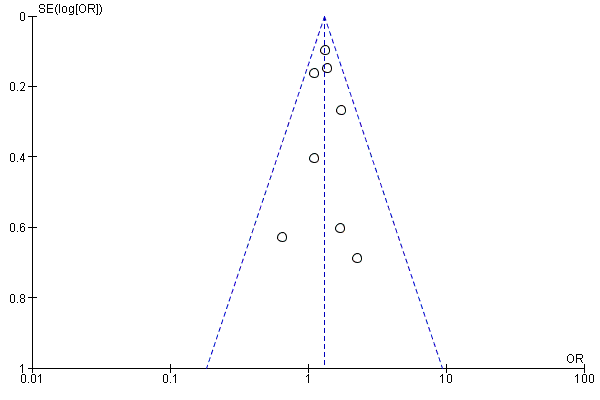

### Funnel plots were generated to judge concerns on publication bias (Supplementary figures).

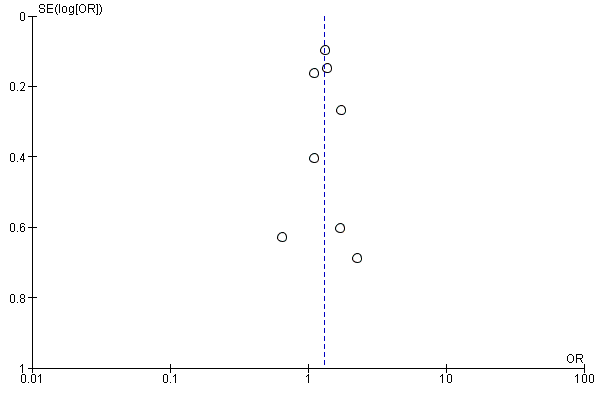

### Funnel plots were generated to judge concerns on publication bias (Supplementary figures).

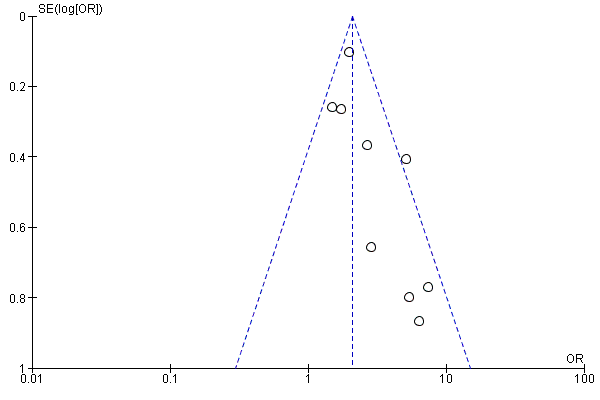

### Funnel plots were generated to judge concerns on publication bias (Supplementary figures).

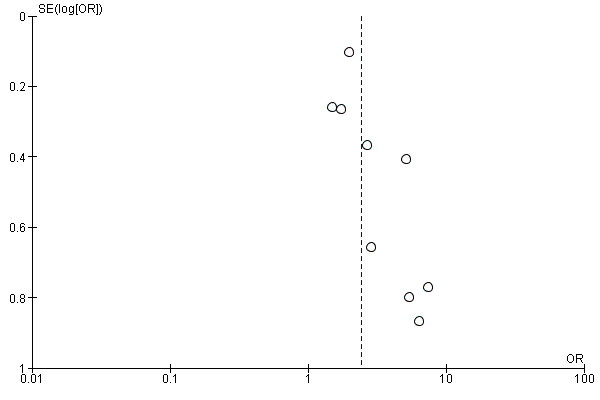
